# Taxane Therapy Alters Expression of BCL2 and Related Protein Partners in White Blood Cells from Breast Cancer Patients

**DOI:** 10.1101/2025.05.15.25327694

**Authors:** S. Munshani, E.Y. Ibrahim, R.L. Rodwin, L.M. Ferrucci, K. Blenman, M. Lustberg, B.E. Ehrlich

## Abstract

The quality of life for many cancer survivors is compromised due to severe, long-lasting side effects of chemotherapy. As part of a preliminary prospective, non-interventional study to examine the side effects of chemotherapy in patients with breast cancer, we examined the change in protein expression levels in blood collected from patients after treatment with taxanes for 12 weeks (n=7). Protein expression levels were measured with reverse-phase proteomic arrays. Here, we examine changes in proteins related to apoptosis, senescence, and calcium signaling in adjuvant vs. neoadjuvant-treated patients The largest change identified was BCL2 (B-cell lymphoma 2), a founding member of the BCL2 family of proteins that regulate apoptosis. Other proteins regulated by BCL2, including RB1 (retinoblastoma protein 1) and NLRP3 (NLR family pyrin domain containing 3) changed significantly over the course of treatment. These differences are relevant to calcium signaling dysregulation and an increased senescent response, both contributors to cancer recurrence. To validate the observations in this small sample, comparisons were made using Kaplan-Meier plots generated from The Cancer Proteome Atlas (TCPA) breast cancer data. The analysis of the TCPA data also shows a large population with upregulation of BCL2 and that elevated BCL2 is associated with a lower survival probability. Once further validated, these findings indicate that the long-term regulation of BCL2 and related proteins should be considered to optimize patient health and prevent recurrence after taxane-based treatment for breast cancer patients.

## Introduction

Taxanes are a family of antineoplastic agents that include paclitaxel and docetaxel. They are drugs widely used as neoadjuvant chemotherapy in several cancer types, including breast, ovarian, and lung cancer [1]. Along with reducing the tumor size, administration of these drugs leads to changes in the expression of a number of proteins [2; 3; 4; 5]. One of the proteins impacted by paclitaxel in breast cancer cells is BCL2, an anti-apoptotic agent that regulates calcium release from the intracellular calcium stores in the endoplasmic reticulum (ER) [6]. BCL2 prevents apoptosis through a complex network that includes the inhibition of RB1 (retinoblastoma protein 1), NLRP3 (NLR family pyrin domain containing 3), and the activation of GRP75 (glucose related protein 75; also known as HSPA9) [7] (Figure 1). RB1 functions to promote cellular senescence and its phosphorylation state is necessary for determining a cell’s fate [8]. NLRP3 is a protein involved in the inflammasome and its activation leads to cell death [9]. GRP75 is a protein that facilitates contact between the ER and mitochondria [10], allowing preferential movement of calcium between these organelles.

**Figure 1.**
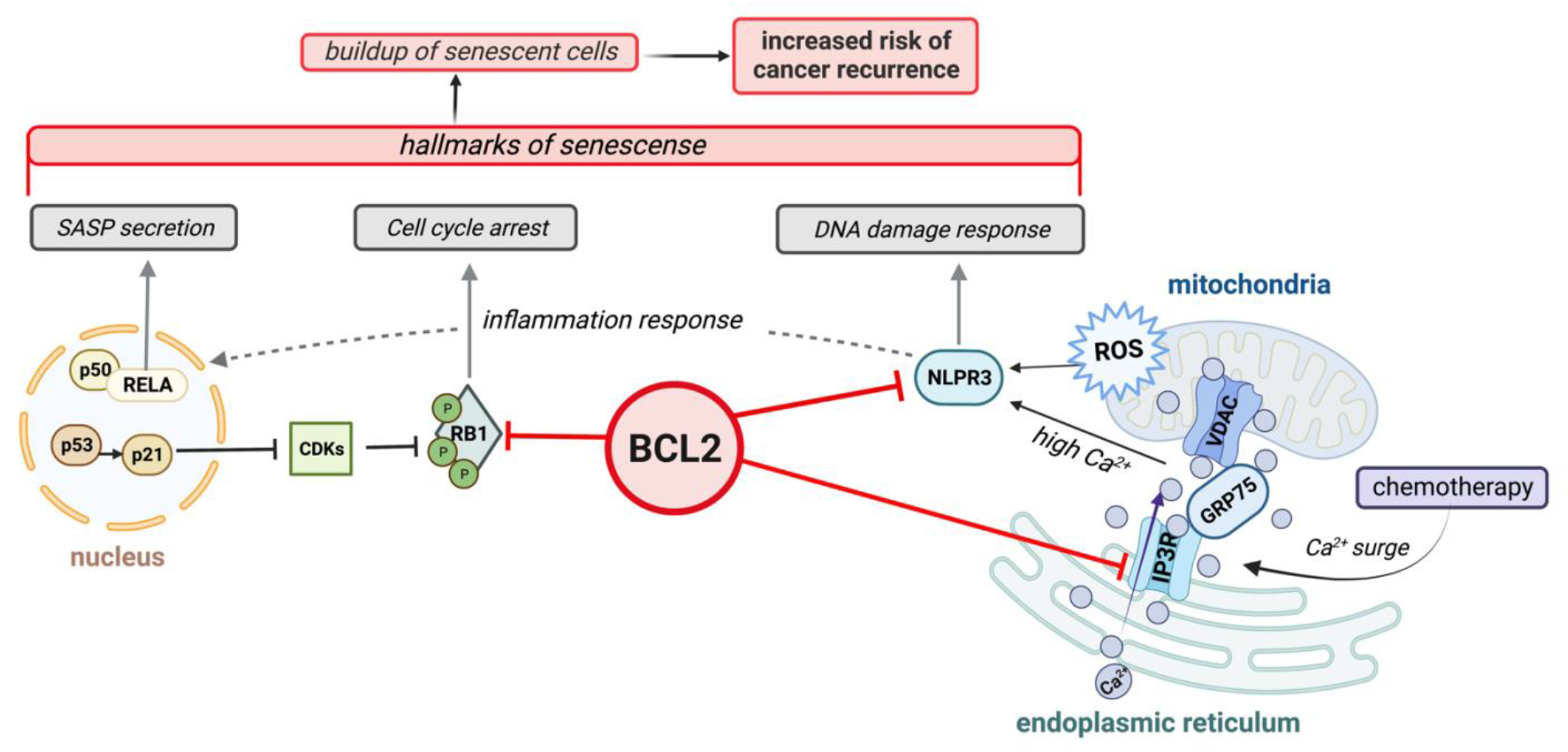
Schematic model showing BCL2 inhibition of key proteins involved in calcium signaling and the senescence pathway.

Calcium signaling dysregulation leads to adverse side effects during taxane-based chemotherapy [11; 12; 13]. In general, calcium signaling is a fundamental process that plays a critical role in cell growth, survival, and death. Alterations in calcium signaling alter many physiological processes which lead to diseases including heart failure, cancer, and neurodegenerative diseases [11; 14; 15]. Relevant to cancer therapy, calcium signaling dysregulation often leads to a state of permanent cell growth arrest known as senescence [16]. The loss of calcium homeostasis is a stress that induces senescence, a state that is the consequence of the changes in the complex interactions between ER and mitochondrial functions [17]. These calcium and stress-induced changes can lead to negative side effects from chemotherapy [18; 19]. These adverse responses include peripheral neuropathy and cognitive impairment [11; 12] and a senescence state where cancer cells become resistant to chemotherapy and radiation [13]. Mitigating the adverse effects of chemotherapy, including neuropathy, senescence, and cognitive impairment, is necessary to improve patients overall health and to avoid treatment holds that can impact survival [20].

In this short report, we examine differences in protein expression of proteins related to apoptosis, senescence, and calcium signaling using samples obtained during a small prospective clinical study that monitored the development of chemotherapy-induced neuropathy in patients with breast cancer who received adjuvant and neoadjuvant taxane-based chemotherapy [21]. Neoadjuvant chemotherapy is a commonly used option for breast cancer patients and it is defined as pharmacological treatment administration prior to surgical intervention [22]. Adjuvant chemotherapies are designed to be delivered after the primary treatment, often surgery, to help to eliminate malignant cells to reduce the chances of recurrence [23]. The largest differences between adjuvant and neoadjuvant groups occurred with proteins related to the pro-apoptotic protein BCL2 and to intracellular calcium signaling. The number of subjects in the prospective study was small, but the results are compatible with conclusions from analysis of large-scale The Cancer Proteome Atlas (TCPA) data. Overall, we examine changes in protein expression levels after 12 weeks of chemotherapy with taxanes in patients with breast cancer to guide further studies into the molecular basis for chemotherapy induced changes in cell function related to cancer treatment and recurrence..

## Methods

We carried out a prospective clinical study of peripheral neuropathy and cognitive function in taxane-treated breast cancer patients with methods previously described [21]. Briefly, studies were approved by the Yale Cancer Center Human Research Ethics Committee (NCT03872141) and written informed consent was obtained in accordance with the Declaration of Helsinki. Patients included in the study were recruited from 08 Aug 2017 until 11 Mar 2020. All data were de-identified. Blood samples were collected in heparinized tubes before the first paclitaxel/docetaxel treatment (baseline) and at 6 weeks and 12 weeks during treatment. White blood cells were isolated, frozen and stored at -80C. To supplement the peripheral neuropathy and cognitive tests reported previously [21], white blood samples were sent to the Reverse Phase Protein Array (RPPA) MD Anderson Core for antibody-based proteomics in March 2021. Seven out of the thirteen patients in the original study provided blood samples at 0 and 12 weeks (baseline and end-of-treatment). Values used for analysis were linearly normalized by MD Anderson, and these were not subsequently manipulated. Week 0 refers to the baseline values before receiving the first doses of paclitaxel/docetaxel. For adjuvant-treated patients, week 0 was around ten weeks after either a partial mastectomy or lumpectomy. The data from these seven patients were used for analysis. A complete list of the proteins analyzed is included in the previous publication [21]. For the analysis, the RPPA data for the seven patients was compared after separation into neoadjuvant (5 patients) and adjuvant (2 patients) treatment with docetaxel and/or paclitaxel.

The Cancer Genome Atlas (TCGA) RPPA BRCA Invasive Carcinoma dataset (downloaded January 2023) was used to analyze protein expression extracted from a large number of patients. TCGA RPPA data provides quantitative protein expression data from validated TCGA cohorts [24; 25; 26]. The TCGA RPPA dataset does not provide an explicit distinction between neoadjuvant, and adjuvant-treated patients [24; 25; 26]. Nonetheless, the analysis provides insight into the impact of the expression of protein levels on survival. In addition, because these data are generated via the MD Anderson core, the sample handling, and data processing parallels well with our patient data. TCPA data and the KMplotter cutoff suggestions were used to generate Kaplan-Meier plots for selected proteins of interest, which were chosen based on the results from our prospective study.

### Limitations

We were limited by the size of the cohort from the prospective study, which was 7 patients. Thus, the patient data and the conclusions are more reminiscent of a case series as opposed to a definitive, large-scale cohort study. In addition, the number of validated proteins included in the RPPA Core is limited to 484 proteins, primarily proteins related to cancer initiation and progression. Therefore, not all the proteins involved in the pathways related to the BCL2 pathway were available for analysis.

## Findings

The patients from the prospective clinical study were divided into two groups: neoadjuvant chemotherapy (n=5) and adjuvant chemotherapy (n=2). The relative change in protein level was calculated from baseline (0 weeks) to end of treatment (12 weeks). Note that baseline levels for neoadjuvant treated patients do not include any pre-treatment data or other metrics prior to the twelve week course of taxane treatment [21]. The expression of the following proteins is highlighted because there was an opposite trend between the neoadjuvant and adjuvant treated patients. The adjuvant treated patients did not receive any chemotherapy prior to the twelve weeks of paclitaxel/docetaxel; instead, they just received either a partial mastectomy or lumpectomy. It should be noted that the adjuvant-treated group did receive a slightly increased dosage of paclitaxel/docetaxel during the twelve weeks, but the treatment regimen for all patients during the twelve weeks was limited to combinations of DDAC and trastuzumab with paclitaxel/docetaxel (Table 1). Additional demographic data is shown in Supplementary Table 4.

**Table 1.** Patient-specific details on HER2/ER/PR status, chemotherapy agents used, weekly dosage of taxol, previous therapies, and weeks post-op (see Supp. Table 4 for demographics).

| Patient | HER2/PR/ER status | Chemotherapy agents | Taxol dosage | Previous therapy | Weeks post- |
| --- | --- | --- | --- | --- | --- |
| *CIPN06* | Triple-positive | Taxol 80x12 then DDAC | Not in chart |  |  |
| *CIPN08* | Triple-negative | Taxol 80x12 then DDAC | 138 mg in 0.9% NaCl |  |  |
| *CIPN09* | HER2+, PR-, ER- | DDAC thenTaxol x12 | Not in chart |  |  |
| *CIPN10* | HER2-, PR+, ER+ | Taxol 80x12 then DDAC | 138 mg in 0.9% NaCl |  |  |
| *CIPN11* | HER2-, PR+, ER+ | DDAC thenTaxol x12 | 156 mg in 0.9% NaCl | Lumpectomy | 13 weeks, 5 days |
| *CIPN12* | Triple-positive | DDAC thenTaxol x12 | 180 mg in 0.9% NaCl | Partial mastectomy | 9 weeks, 3 d |
| *CIPN13* | HER2-, PR+, ER+ | Taxol 80x12 then DDAC | 138 mg in 0.9% NaCl |  |  |

### Main Focus: BCL2’s relationship with NLRP3, GRP75, and RB1

After dividing the patients into adjuvant and neoadjuvant treatment groups, data were sorted to identify proteins with a greater than statistically significant difference in the relative change in protein level between the two treatment groups. Of the 484 proteins evaluated by RPPA, the protein with the largest difference between neoadjuvant and adjuvant treated patients was BCL2 (Figure 2A). A list of the proteins with the largest differences shows a bias toward proteins in the BCL2 family, calcium signaling complex (Figure 2B), and senescence pathway (Figure 2C). Of the top hits, neoadjuvant and adjuvant treated patients showed a distinct and opposite trend in protein expression for BCL2, NLRP3, and RB1. For adjuvant treated patients, BCL2 decreased by 37% (q1: -47%, q3: -28%), NLRP3 increased by 68% (q1: 51%, q3: 84%), and RB1 increased by 15% (q1: 6%, q3: 21%). For neoadjuvant treated patients, BCL2 increased by 67% (q1: 39%, q3: 103%), NLRP3 decreased by 8% (q1: -19%, q3: 3%), and RB1 decreased by 12% (q1: 0%, q3: -15%) (Supplementary Table 3). These results show that BCL2 had the most significant difference between adjuvant and neoadjuvant treated patients (Supplementary Figure 1, Supplementary Table 2). Previous reports indicate that paclitaxel treatment of cancer cell cultures lead to phosphorylation of BCL2 [27], a modification that determines a cell’s fate [8]. The changes reported here in cells from human subjects validate the previous cultured cell-based work. Note that BCL2 also can prevent apoptosis via inhibition of NLRP3 and RB1.

**Figure 2.**
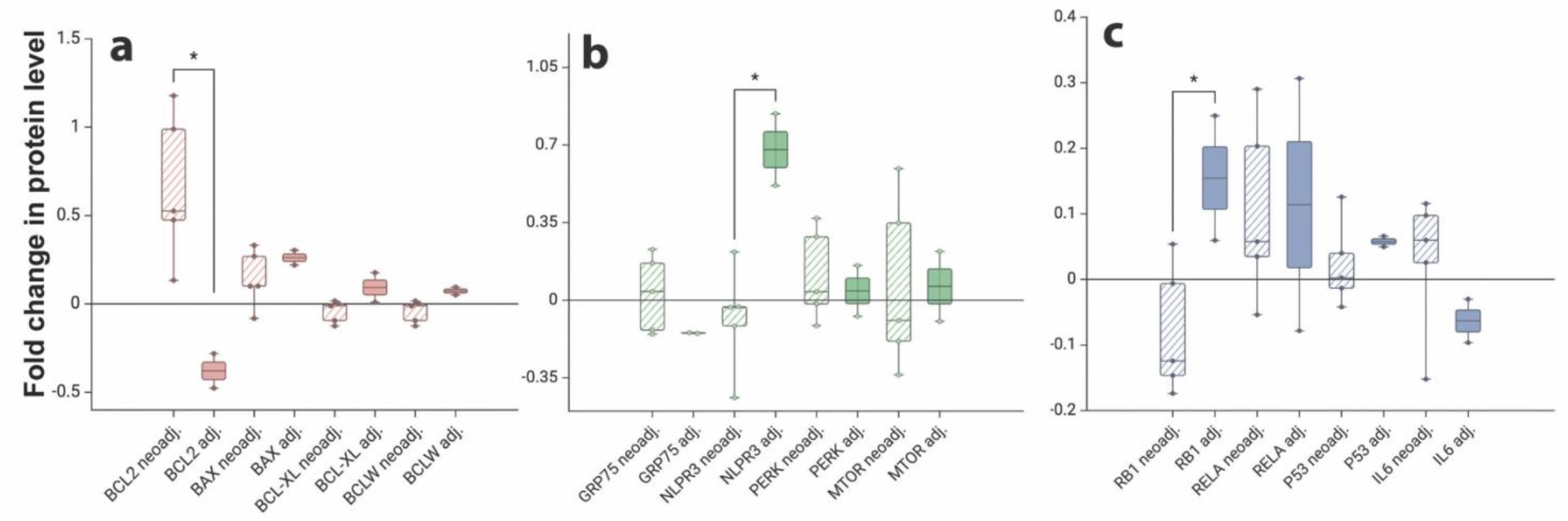
A-C: Relative change in protein expression for neoadjuvant (n=5) and adjuvant (n=2) patients over the course of 12 weeks of treatment. Figures 2a-2c demonstrate proteins with notable changes within the BCL2 family (Fig. 2a), calcium signaling associated proteins (Fig. 2b), and senescence associated proteins (Fig. 2c). For proteins with a significant difference (see Supp. Table 2 for unpaired t-test, Supp. Table 3 for descriptive statistics) in expression after treatment between neoadjuvant and adjuvant groups.

### The BCL2 Family

Given that adjuvant and neoadjuvant patients showed the greatest divergence in BCL2 protein level (Figure 2A), we analyzed the available BCL2 family members in the patient RPPA data. BAX, BCL-XL, BCLW, BIM showed small differences between the two cohorts (Figure 2A). For all of these members of the BCL2 family except BIM and PUMA (not shown), the neoadjuvant patients had a slight decrease in protein levels relative to the adjuvant patients (Supplementary Table 1).

BCL2 is central to anti-apoptotic processes through its ability to interfere with the actions of pro-apoptotic proteins, BCL-XL, BAX, BCLW, and BIM. BCL-XL is directly involved with mitochondrial homeostasis, and has been reported to prevent apoptosis and control breast cancer cell migration via mitochondrial reactive oxygen species (ROS) formation [28]. Furthermore, it has been shown that Nrf2 binds to BCL-2 gene antioxidant response element to control both BCL2 activity and cellular apoptosis [29], and also promotes chemoresistance [30]. In general, adjuvant patients had an increase in the expression levels of these pro-apoptotic proteins whereas neoadjuvant patients had a decrease in expression levels, suggesting that pro-apoptotic behavior is downregulated when BCL2 levels increases. BCL2 also impacts the behavior of the inositol tris-phosphate receptor (ITPR) where BCL2 binds to and inhibits the calcium channel activity of the intracellular calcium channel formed by ITPR [31; 32]. In addition to the role of BCL2 in down-regulating the activity of ITPR3, other studies report that BCL-XL sensitizes the ITPR to low levels of inositol tris-phosphate, the agonist of ITPR channels, and that BCL-XL binds to the same region of the ITPR as BCL2 [33; 34]. The potential competition between BCL2 and BCL-XL emphasizes the importance of the relative expression of these two proteins. Once bound to the ITPR at the mitochondrial-associated ER membrane (MAM), BCL-XL promotes *pro-survival* oscillations of calcium ions whereas BCL2 prevents *pro-apoptotic* transients of calcium ions [35; 36].

### Calcium Signaling

Proteins that were involved in calcium signaling, particularly at the MAM, showed divergence between adjuvant and neoadjuvant treated patients (Figure 2B). As previously mentioned, BCL2 influences calcium signaling by inhibiting the ITPR. Due to the limits of the RPPA patient data, we could not evaluate all relevant players involved in calcium signaling, but it was possible to evaluate PERK, GRP75, and MTOR alongside BCL2 and NLRP3 (Figure 2B). PERK, GRP75, and MTOR levels did not deviate dramatically between adjuvant and neoadjuvant treated patients. As these proteins are cytoplasmic or plasma membrane associated, this result suggests that the relevant signaling pathway is more focused to the MAM, particularly the calcium ion regulation mitigated by ITPR channel activity.

NLRP3 is a calcium-mediated protein associated with the inflammasome, whose activation is triggered by mitochondrial ROS production and by ER stress [37]. Once NLRP3 is activated, NLRP3 is known to trigger various types of cell death including pyroptosis, necrosis, and apoptosis [38]. Furthermore, BCL2 and BCL2-XL suppress NLRP3 activity by binding ATP and preventing oligomerization of NLRP3 [37; 39]. Calcium activation is necessary for the NLRP3-dependent pro-inflammatory action to occur, which is partially mediated by the GRP75-VDAC1-ITPR3 complex [40; 41]. GRP75 is a glucose-regulated protein that facilitates contact between the ER and mitochondria via the MAM and is a tether protein that links the ITPR and voltage dependent anion channel (VDAC1) to regulate calcium flux [42; 43]. BCL2 interferes in this pathway due to its binding to the ITPR which will inhibit the release of calcium from the ER and the transfer of calcium to the mitochondria [44; 45].

### Cellular Senescence

RB1 is known to be involved in senescence, and RB1 dephosphorylation plays a role in determining a cell’s apoptotic fate [46; 47]. When RB1 is inhibited (i.e., not dephosphorylated), the cell does not undergo apoptosis. Specifically, previous studies have indicated that BCL2 anti-apoptotic activity prevents RB1 dephosphorylation and apoptosis thereby allowing RB1 to activate senescence related pathways [48]. Additionally, BCL2 has been shown to modulate Hippo signaling [49]. The Hippo signaling pathway has been shown to be involved in creating resistance to therapeutics [50] and the induction of senescence in human nucleus pulposus chondrocytes leads to an increase of the Hippo pathway [51].

Recent evidence demonstrates that the primary pathways underlying senescence are calcium signaling related [52]. Cancer treatments are known to result in cellular senescence, which occurs when the cell cycle is halted and unable to repair damage. However, in some cases, the cell never restarts division nor does apoptosis occur, yet the cell remains in a senescent state marked by the secretion of SASP (senescent associated secretory phenotypes) [52]. SASP production encourages an environment that is immunosuppressive and increases the proliferation of surrounding cells, which results in a recurrence of cancer [13; 52; 53; 54]. During senescence, the interaction between ITPR and the MAM increases and there is an accumulation of calcium ions in the mitochondria [42; 55]. Due to the altered mitochondrial state, ROS production increases. The increased ROS often leads to DNA damage, which triggers DNA damage response (DDR) pathways, and activates p21 to inhibit cyclin-dependent kinases. The activation of these kinases then inhibits pro-apoptotic protein RB1, leading to secretion of SASP proteins, such as IL6 [56; 57; 58].

Only one of the documented SASP proteins, IL6, an interleukin, was available within the RPPA patient dataset. IL6 expression levels in adjuvant patients was lower after 12 weeks to chemotherapy, whereas neoadjuvant patients showed higher expression of IL6 (Figure 2C). Additional senescence-related proteins were examined: RELA, which is an anti-apoptotic protein involved in coordinating the DNA damage response (DDR) and SASP secretion, and p53, which is a tumor suppressor that promotes apoptosis [59; 60; 61]. P53 also inhibits Nf-kB, which activates BCL2. Our results indicate that p53 and RELA levels were similar for neoadjuvant and adjuvant patients, possibly suggesting that p53 was unable to regulate BCL2 activity via Nf-kB. These comparisons further indicate the possibility that neoadjuvant patients have a higher incidence of senescent cells.

### Long-Term Implications

Our patient data was limited to the scope of the prospective study, which was the length of taxane treatment (12 weeks). However, the potential implication of these findings is better contextualized using longitudinal data. To investigate long-term implications, Kaplan-Meier plots were generated using open access RPPA data. Despite low patient numbers for NLRP3, RPPA data was used instead of gene chip or mRNA data to remain consistent with the usage of proteomics data in this report. The open access data were not split by neoadjuvant and adjuvant patients, which means these data reflect general trends across a breast cancer cohort (n=875 for BCL2 and RB1, n=65 for NLRP3). The data were split by optimal cutoff based on ROC curves, and overall survival (OS) data was plotted over the course of 120 months (Figure 3). KMplotter, a verified Kaplan-Meier curve generator was used to visualize the curves [62]. The proteins examined for the Kaplan-Meier plots were BCL2 (Figure 3A), NLRP3 (Figure 3B), and RB1 (Figure 3C). For NLRP3, only data from an external study [63] was available, whereas for BCL2 and RB1, data from the TCGA RPPA was available. All three of these proteins showed the same relative pattern. For BCL2 and RB1 the high and low expression level groups were mostly in line with one another until approximately the 2.5-year mark. After this point, patients who had high expression of BCL2 and RB1, had a lower probability of survival. In contrast, for NLRP3, the divergence occurred later. Considering that all neoadjuvant patients in the Kaplan-Meier plots exhibited BCL2 overexpression right after completing treatment, these results suggest that regulation of BCL2 levels is integral to overall survival.

**Figure 3.**
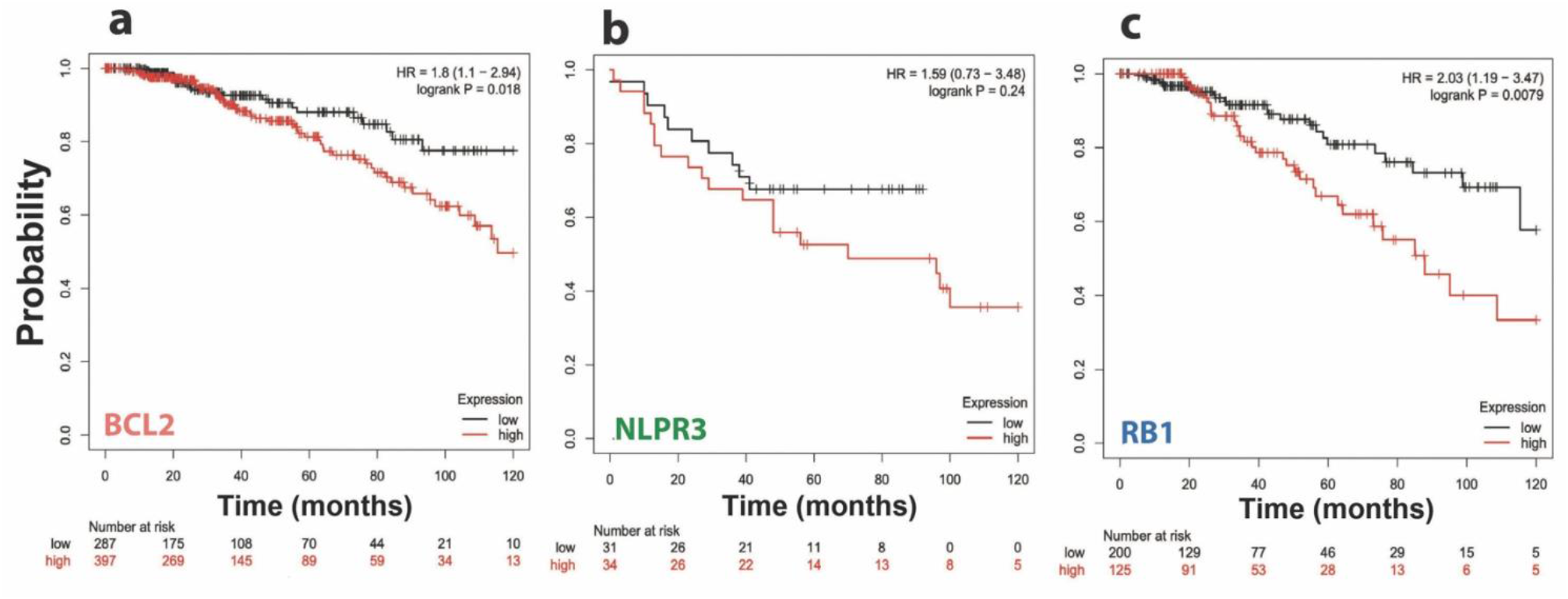
A-C: Kaplan-Meier survival plots for proteins identified in Figure 2. Survival plots are over the course of 120 months for BCL2 (HR = 1.8 (1.1-2.94), logrank P = 0.018) (panel A), NLRP3 (HR = 1.59 (0.73-3.48), logrank P = 0.24) (panel B), and RB1 (HR = 2.03 (1.19 – 3.47), logrank P = 0.079) (panel C). Survival cutoff was set to 120 months and patients were split by optimal cutoff. (Note: red = overexpression, black = underexpression).

It is worth noting that there is a distinction between long-term and short-term effects of taxane-based treatment. Shortly after treatment, BCL2 shows an immediate increase in expression in the majority of patients (neoadjuvant). Additionally, although the short-term effects NLRP3 and RB1 are less dramatic, the Kaplan-Meier plots demonstrate adverse long term-effects associated with the overexpression of these proteins. The increase of all these proteins implicates long-term effects in the senescent pathway, such that the buildup of senescent cells post-taxane treatment is a result that becomes apparent months after treatment, as opposed to just weeks after treatment.

### Discussion /Conclusion

The purpose of this study was to investigate BCL2 and associated protein expression levels in a cohort of patients with breast cancer who underwent adjuvant or neoadjuvant taxane-based chemotherapy. Overall, we found that breast cancer patients who received neoadjuvant chemotherapy had significantly increased levels of the protein BCL2 compared to those who received adjuvant chemotherapy. Other proteins related to BCL2, NLRP3, and RB1, also showed differences between the two groups. These differences may contribute to calcium signaling dysregulation and an increased senescent response from cells, both of which can increase the risk of cancer recurrence. In addition, within the larger dataset, upregulation of BCL2 and related proteins was associated with a lower survival probability 2-3 years after treatment. However, it is important to note that the study was limited by its small size and the limited number of validated proteins available for analysis. Further research with larger sample sizes and a wider range of proteins is needed to confirm and expand upon these findings. If validated in larger datasets, we suggest that when considering neoadjuvant and adjuvant treatment, along with the long-term implications of chemotherapy, it is important to also consider calcium signaling dysregulation and the potential for chronic senescence. The goal of cancer treatment is not only to eliminate the immediate threat, but also to prevent recurrence. Thus, regulating BCL2 activity is one possible route to maintain long-term cellular homeostasis and prevent recurrence of cancer metastasis.

## Data Availability

All datasets used were either publicly available or have been included in previous publication https://doi.org/10.1371/journal.pone.0275648

## Table and Figure Legends

Supplementary Figure 1: Comparison and significance of changes in BCL2, RB1, NLPR3 levels from 0-12 weeks of treatment in neoadjuvant (n=5) and adjuvant (n=2) treated patients. Unpaired t-test was performed to determine statistical significance of differences.

Supplementary Table 1: Percent change in protein expression levels from 0 to 12 weeks after taxane-based treatment for neoadjuvant (n=5) and adjuvant (n=2) patients.

Supplementary Table 2: Unpaired t-test between adjuvant-treated and neoadjuvant-treated patient groups for proteins with significant deviations (p<0.05).

Supplementary Table 3: Descriptive statistics for proteins with p<0.05.

Supplementary Table 4: Patient demographics. Age and BMI are reported as Mean +/-SD.

## Statements and Declarations

### Author contributions

SM analyzed data and wrote the first draft; EYI obtained and analyzed data, edited manuscript; RLR added clinical insights and edited manuscript; LMF discussed and edited manuscript; KB discussed and edited manuscript; ML obtained funding, added clinical insights, and edited manuscript; BEE conceived project, obtained funding, analyzed data, and edited manuscript. All authors approved final manuscript.

### Ethics approval

Studies were approved by the Yale Cancer Center Human Research Ethics Committee (NCT03872141) and written informed consent was obtained in accordance with the Declaration of Helsinki.

### Consent to participate

Informed consent was obtained from all individual participants included in the study.

### Consent to publish

The authors affirm that human research participants provided informed consent for the publication of related data.

