## Supplemental Information for "Taxane Therapy Alters Expression of BCL2 and Related Protein Partners in White Blood Cells from Breast Cancer Patients"

**Supplementary figure and tables**

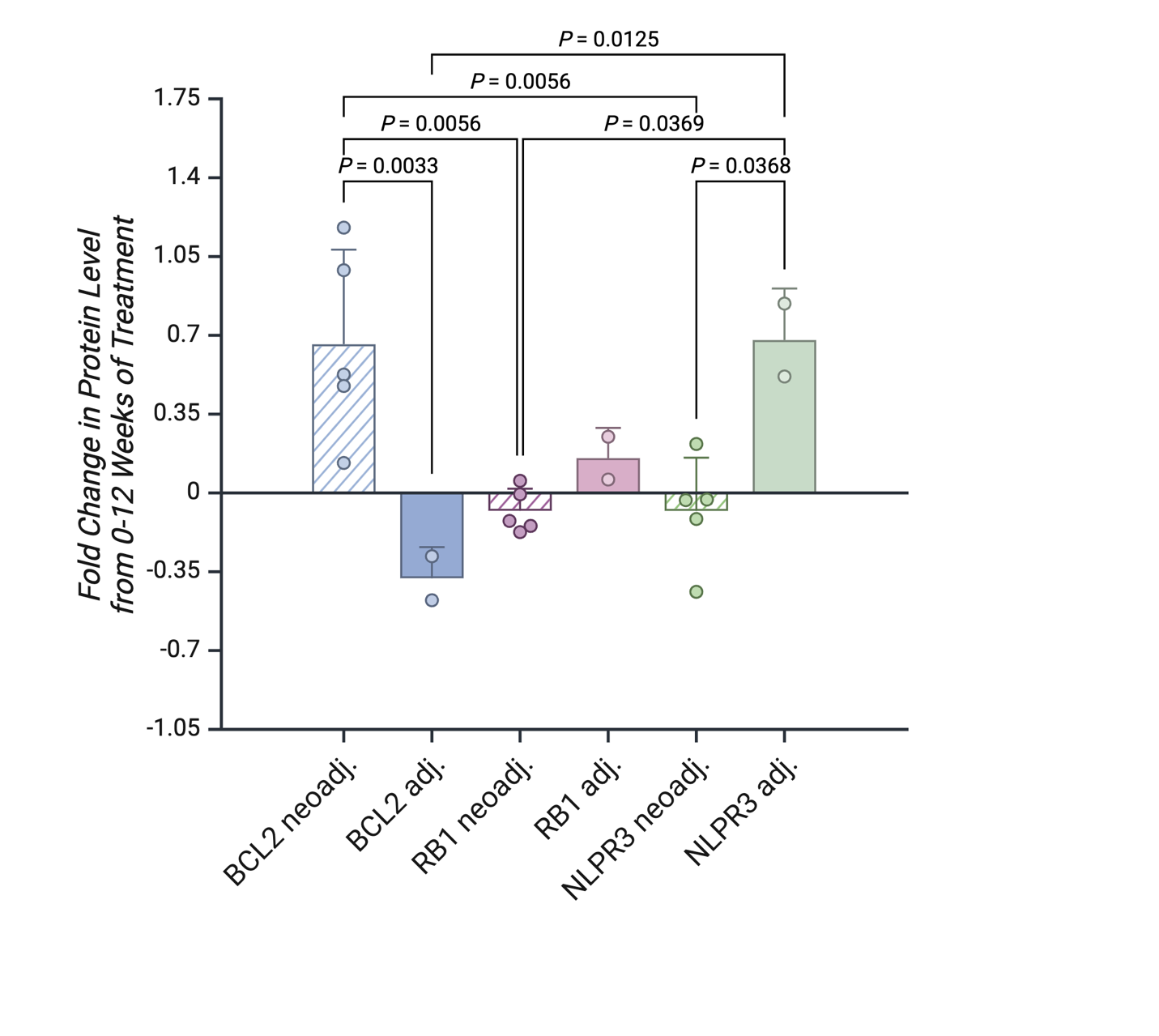

Supplementary Figure 1: Comparison and significance of changes in BCL2, RB1, NLPR3 levels from 0-12 weeks of treatment in neoadjuvant (n=5) and adjuvant (n=2) treated patients. Unpaired t-test was performed to determine statistical significance of differences.

| Protein | Neoadjuvant | Adjuvant |
| --- | --- | --- |
| BCL2 | +67% | –38% |
| NLPR3 | –8.0% | +68% |
| RB1 | +6.4% | +15% |
| GRP75 | +17% | –14% |
| RELA | +10% | +11% |
| P53 | +2.2% | +5.8% |
| IL6 | +11% | – 4.1% |
| PERK | +11% | +4.2% |
| MTOR | +6.5% | +6.2% |
| BAX | +14% | +26% |
| BCL-XL | +5.9% | +9.3% |
| BCLW | –4.2% | +7.3% |

Supp. Table 1: Percent change in protein expression levels from 0 to 12 weeks after taxane-based treatment. neoadjuvant (n=5) and adjuvant (n=2) patients*.*

|  | *t* | *DF* | *p value (two-tailed)* | *Difference Between Means (A - B) ± SEM* | *95% Confidence Interval* | *Cohen's d* |
| --- | --- | --- | --- | --- | --- | --- |
| *BCL2* | 3.2598 | 5 | 0.0225 | -1.0390 ± 0.3187 | 0.2197 to 1.8584 | -2.7273 |
| *RB1* | -2.6285 | 5 | 0.0466 | 0.2331 ± 0.0890 | -0.4628 to -0.0052 | 2.1991 |
| *NLRP3* | -3.86 | 5 | 0.0119 | 0.7588 ± 0.1966 | -1.2640 to -0.2535 | 3.2295 |

Supplementary Table 2: Unpaired t-test between adjuvant-treated and neoadjuvant-treated patient groups for proteins with significant deviations (p<0.05).

|  | *Mean* | *SD* | *SEM* | *Minimum* | *25^th^ percentile* | *Median* | *75^th^ percentile* | *Maximum* |
| --- | --- | --- | --- | --- | --- | --- | --- | --- |
| *BCL2: Neoadjuvant* | 0.6602 | 0.4203 | 0.1871 | 0.1332 | 0.3040 | 0.5256 | 1.0836 | 1.1782 |
| *BCL2: Adjuvant* | -0.3789 | 0.1382 | 0.0977 | -0.4766 | -0.4766 | -0.3789 | -0.2811 | -0.2811 |
| *RB1: Neoadjuvant* | -0.0794 | 0.0982 | 0.0439 | -0.1739 | -0.1602 | -0.1242 | 0.0238 | 0.0538 |
| *RB1: Adjuvant* | 0.1546 | 0.1344 | 0.0950 | 0.0595 | 0.0595 | 0.1546 | 0.2496 | 0.2496 |
| *NLRP3: Neoadjuvant* | -0.0798 | 0.2364 | 0.1057 | -0.4932 | -0.2774 | -0.0321 | 0.0940 | 0.2174 |
| *NLRP3: Adjuvant* | 0.6781 | 0.2281 | 0.1619 | 0.5170 | 0.5170 | 0.6781 | 0.8409 | 0.8409 |

Supplementary Table 3: Descriptive statistics for proteins with p<0.05.

| *Age* | 47 *±*11.7 |
| --- | --- |
| *BMI* | 28.4 *±* 8.6 |
| *Stage* | 1A (n=2)  2A (n=2)  2B (n=2)  N/A (n=1) |
| *History of Smoking* | Yes (n=2)  No (n=5) |
| *Race* | White (n=5)  Black (n=1)  Hispanic (n=1) |

Supplementary Table 4: Patient demographics. Age and BMI are reported as Mean *±* SD.
